# Understanding the relationship between sleep disturbance and allostatic load: A cohort study of women with and without breast cancer

**DOI:** 10.1101/2025.11.18.25340501

**Authors:** Ian R. Nataro, Yufan Guan, Philip I. Chow, Hua Zhao

## Abstract

Breast cancer is one of the most commonly diagnosed forms of cancer, and recent advances in treatment have led to increasing prevalence of breast cancer survivors globally. But even after completing active treatment, breast cancer survivors remain vulnerable to a range of adverse health outcomes due to high levels of physiological stress. This study investigates the relationship between sleep disturbance and allostatic load (AL) in breast cancer survivors and control subjects, using a large cohort sample drawn from the UK Biobank. Breast cancer survivors exhibited a higher mean AL at baseline (m = 2.74) compared to the control sample (m = 2.44; *p* < 0.01), and AL remained elevated as time elapsed following cancer diagnosis. There was a positive association at baseline between AL and symptoms of sleep disturbance in both breast cancer survivors and controls. However, cancer survivors exhibited a greater increase in levels of AL with only moderate or occasional sleep disturbance. Longitudinal analyses revealed that for both cancer survivors and controls, higher AL at baseline predicted poorer sleep quality at the next wave (r ≈ .22; p < 0.05). These findings support a bidirectional relationship between sleep disturbance and overall physiological stress, highlighting sleep as a potential target for intervention in breast cancer survivorship care.

## Introduction

Chronic sleep disturbance, including disorders like insomnia, increases risk for a wide range of adverse health outcomes (Scott et al., 2021; Carroll et al., 2015; Briançon-Marjollet et al., 2015). While insomnia in the general population has been acknowledged as a public health issue, the relationship between sleep and health in cancer survivors has received less attention. This is critical because sleep disturbance is a prominent concern among cancer survivors, with about half reporting meaningful sleep problems (Emery et al., 2022) that often persist after treatment ends (Lowery-Allison et al., 2018). The prevalence of sleep disturbance is markedly high among breast cancer survivors, with estimates up to 85% (Wu et al., 2023). Breast cancer survivors are particularly susceptible to chronic sleep disturbances given their confluence of risk factors including female sex (Jaussent et al., 2011), hormonal therapies, and menopause (Leysen et al., 2019). Sleep disturbance in breast cancer survivors is associated with poor metabolic regulation, greater inflammation (Palesh et al., 2013), low quality of life (Fortner et al., 2002), and high psychological distress (Lowery-Allison et al., 2018), which highlights the urgent need to better understand the relationship between sleep and overall health in this population.

Adequate sleep is especially important for breast cancer survivors because it mitigates the negative impacts of stress and promotes resilience (van Dalfsen & Markus, 2018). Allostatic load (AL) is a composite score reflecting the cumulative physiological burden incurred through exposure to chronic environmental stressors (Juster et al., 2016). AL reflects the ‘wear and tear’ on the body that results from adapting to stress through allostasis, or the process of continual adjustment to maintain physiological stability in unpredictable conditions. Allostasis is mediated in large part by the hypothalamic-pituitary-adrenal (HPA) axis; under conditions of repeated high stress the HPA axis may become dysregulated, leading to an overactive or blunted cortisol response (van Dalfsen & Markus, 2018) and metabolic dysfunction (Kivimaki et al., 2022). Chronically elevated levels of AL are associated with stroke, cardiovascular disease, diabetes mellitus, and all-cause mortality (Akinyemiju et al., 2020). Previous studies have found that breast cancer survivors have higher AL than the general population (Guan et al., 2023), making them vulnerable to adverse health outcomes. It is therefore critical to understand the causes and correlates of high AL in breast cancer survivors to inform targeted behavioral interventions.

Previous studies have uncovered an association between sleep and AL (Clark et al., 2014; Carroll et al., 2015) and increasingly, researchers have posited a bidirectional relationship that can create a vicious cycle, where worsening in one domain contributes to deterioration in the other (Christensen et al., 2022). Sleep disturbance itself functions as a chronic stressor that can alter immune and inflammatory processes, compounding disease vulnerability (Faraut et al., 2012). Likewise, high AL may contribute to conditions such obstructive sleep apnea, vascular disease, and musculoskeletal pain that tend to disrupt sleep (Figorilli et al., 2025). To date, no studies have been published on the relationship between sleep and AL in breast cancer survivors. Therefore, the primary goal of this study is to characterize the relationship between sleep disturbance and AL in a large sample of breast cancer survivors and controls from the UK BioBank data repository. We hypothesized that at baseline, women with a history of breast cancer would evidence both higher AL and symptoms of sleep disturbance (nighttime insomnia and daytime sleepiness) than women with no cancer history. We further hypothesized that higher AL at baseline would predict poorer sleep quality at the next follow-up wave, for both breast cancer survivors and controls.

## Methods

### Study Cohort

We utilized data from the UK Biobank, a nationwide prospective cohort that enrolled over 500,000 UK residents aged 46 to 69 years between 2006 and 2010. At enrollment, participants provided detailed information through touchscreen questionnaires, contributed biological samples, and underwent physical measurements (Sudlow et al., 2015). Additional information about the UK Biobank can be accessed at http://www.ukbiobank.ac.uk/.

This analysis included 177,429 women (defined as female sex reported in database), 5,491 of whom were identified as breast cancer survivors. Participant sex was ascertained by UK Biobank based on self-report and available medical records. Health history was based on self-report medical history gathered by a medical professional during study intake. Individuals with a history of other cancer types were excluded from this sample. Racial and ethnic makeup of the sample were representative of the U.K. population from which it was drawn. Biobank participants missing any of the eleven biomarkers needed to compute AL were also excluded. Time interval between intake and follow-up data collection incidents varies by participant, with a reported median follow-up interval of 12 years (Allen et al., 2024). The sample size was determined by the availability of eligible participants in the database. Because this study used existing data from the UK Biobank, no a priori power calculation was performed. All participants meeting inclusion criteria were included to maximize statistical power and representativeness. This study was approved by the institutional review board at University of Virginia.

### AL Score Construction

Using an established method, AL score was constructed from eleven baseline biomarkers across cardiovascular, inflammatory, and metabolic systems (Shen et al., 2022; Guan et al., 2023). The cardiovascular domain included systolic blood pressure (SBP), diastolic blood pressure (DBP), and pulse rate (PR); the inflammatory domain C-reactive protein (CRP); and the metabolic domain high-density lipoprotein (HDL) cholesterol, waist-to-hip ratio, total cholesterol, triglycerides (TG), hemoglobin A1c (HbA1c), and creatinine. Each biomarker was classified into high- or low-risk categories based on established clinical cutoffs, with high-risk values assigned a score of one (Zhao et al., 2021). Participants reporting the use of medications for hypertension or metabolic disorders were also assigned a score of one in the corresponding category. AL score was calculated as the sum of all high-risk scores, with a maximum possible score of 11. Higher AL scores reflect greater physiological stress.

### Sleep Disturbance

Baseline sleep disturbance was evaluated through self-report measures concerning insomnia symptoms and daytime sleepiness. Insomnia was evaluated with the question: “Do you have trouble falling asleep at night or do you wake up in the middle of the night?” Response options included: never/rarely, sometimes, and usually. Daytime sleepiness was assessed by asking, “How likely are you to doze off or fall asleep during the daytime when you don’t intend to? (e.g., while working, reading, or driving),” with response options: never/rarely, sometimes, and usually. An assessment of sleep quality at the next wave was administered using the Pittsburgh Sleep Quality Index (PSQI), a validated instrument that captures multiple dimensions of sleep including measures of insomnia (Niu et al., 2023). Detailed methodology regarding the PSQI has been previously described by Buysse et al. (1989).

### Assessment of Covariates

To strengthen the validity of this analysis, we adjusted for a broad range of covariates encompassing demographic, socioeconomic, and lifestyle factors. Demographic information included age and race. Socioeconomic variables comprised education, household income, and Townsend Deprivation Index (TDI). TDI is a geographic measure of material deprivation based on residential postcode. Income was dichotomized at £31,000, and TDI was categorized as high or low using the cohort median. Educational attainment was classified as either “High school or less” or “College/professional.” Lifestyle factors included cigarette smoking, alcohol consumption, and physical activity. Smoking status was derived from self-reported current and past use of inhaled tobacco. Alcohol intake frequency was categorized as “special occasions or never,” “moderate”, or “heavy”. Physical activity was evaluated using Metabolic Equivalent of Task (MET) scores and categorized as low, moderate, or high.

### Statistical Analysis

We employed a multistep analytical approach to examine the associations between allostatic load (AL), insomnia and daytime sleepiness among breast cancer survivors and a control sample of women with no cancer history. Baseline demographic and clinical characteristics were compared between the two groups using independent t-tests for continuous variables and chi-square tests for categorical variables. AL scores were analyzed both as continuous and categorical variables to assess distributional differences between groups (Supplemental Figure 1; Supplemental Table 2). To examine differences in allostatic load (AL) by time since breast cancer diagnosis, we generated boxplots (Figure 1) stratifying the case sample into five groups: ≤ 2 years, 2–5 years, 5–10 years, and >10 years since diagnosis. Mean AL values were overlaid for all strata and controls; p-trend was calculated to assess linear associations across these ordinal categories. To evaluate the association between sleep behaviors and AL, linear regression models were fitted separately for breast cancer survivors and non-cancer women. Insomnia and daytime sleepiness were each categorized into three degrees of severity based on symptom frequency. Two regression models were constructed: an unadjusted (crude) model and a multivariable-adjusted model controlling for potential confounds. These included age at recruitment, education level, household income, TDI, smoking status, alcohol consumption, and physical activity (see next section on sample characteristics). Trends across categories of symptom severity were examined within each group, and interaction terms were used to test for differential effects between groups. Adjusted means and standard errors were reported. We further explored whether baseline AL predicted subsequent sleep problems assessed through the Pittsburgh Sleep Quality Index (PSQI) and its subcomponents. Separate linear regression analyses were conducted for each sleep domain, with AL as the primary independent variable. Models were stratified by breast cancer status and adjusted for the same covariates as above. Results were expressed as regression coefficients with 95% confidence intervals. All statistical tests were two-sided, and a p-value < 0.05 was considered statistically significant.

**Figure 1.**
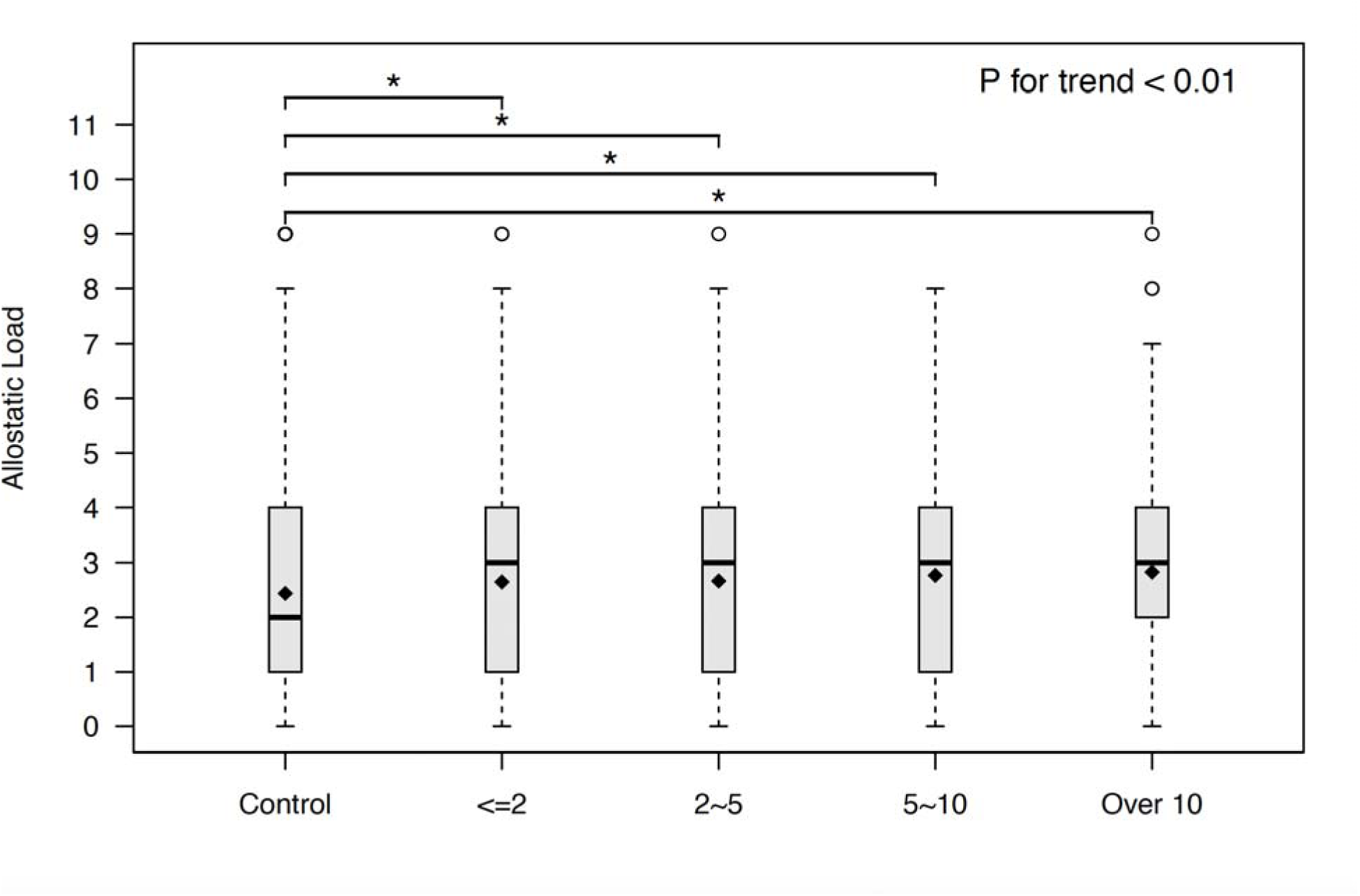
Allostatic load based on time since cancer diagnosis.

Analyses were conducted using R version 4.3.0.

## Results

### Characteristics of Sample

Supplemental Table 1 summarizes selected demographic and lifestyle characteristics of the full sample. Results of a chi-square test found that the breast cancer and non-cancer (control) samples differed significantly in age, racial composition, income, smoking status, and physical activity (*p* ≤ 0.01) Breast cancer survivors were an average of 3.5 years older than controls and had a 4.8% higher rate of past or present smoking. They were 5.4% more likely to report an annual family income below £31,000. Physical activity was overall lower among the breast cancer sample, but this comparison was limited by missing data. The frequency of insomnia symptoms among the breast cancer sample aligns with existing research (Wu et al., 2023; Leysen et al., 2019), with 38% of the sample reporting usual insomnia compared to 31.8% of the control sample. Breast cancer survivors were also more likely to report daytime sleepiness, with an additional 3.8% endorsing this symptom to some extent compared to controls. Chi-square tests performed on sleep disturbance symptoms were both significant at *p* < 0.01.

### Distribution of Allostatic Load

A chi-square test indicated a significant group difference in the distribution of AL scores between the breast cancer (case) and control samples (*p* < 0.01). Breast cancer survivors had a higher mean AL (μ = 2.74) than the control sample (μ = 2.44). These results can be viewed in Supplemental Figure 1 and Supplemental Table 2. The distribution of AL scores in the breast cancer sample is positively skewed to a greater degree than the distribution of scores of the case sample, indicating that AL is more normally distributed among breast cancer survivors vs. controls, which likely accounts for their higher mean AL. Figure 1 shows the distribution of AL scores in breast cancer survivors compared to controls, with the breast cancer sample stratified by years since cancer diagnosis at the time of study recruitment, given that those more recently diagnosed may face greater stressors than those diagnosed longer ago. A significant increase in AL was observed over time since diagnosis (*p* < 0.01), indicating that as time elapses following a breast cancer diagnosis, AL does not stabilize or regress to the control sample mean.

### Relationship between Allostatic Load, Insomnia and Daytime Sleepiness

Table 1 compares mean AL scores in the breast cancer and control groups stratified by severity of insomnia and daytime sleepiness in both crude and adjusted models. In the crude model, before controlling for covariates, there were significant within-group differences for both breast cancer survivors and controls, such that having insomnia or daytime sleepiness ‘sometime’ or ‘usually’ was significantly associated with higher AL when compared to no insomnia/daytime sleepiness (all *p’s* < 0.01). Moreover, there were significant between-group differences such that breast cancer survivors reporting having insomnia ‘sometime’ or ‘usually’ had higher AL compared to controls reporting the same severity of insomnia symptoms (*p* < 0.01). Breast cancer survivors reporting having daytime sleepiness ‘sometime’ also had higher AL compared to controls reporting the same severity of daytime sleepiness symptoms (*p* < 0.01).

**Table 1.** Levels of allostatic load by degree of insomnia and daytime sleepiness in breast cancer survivors and controls.

|  | Non-cancer<br>wome | P value<sup>#</sup> | Crude model<br>Breast cancer<br>survivors | P value<sup>#</sup> | P value<sup>@</sup> | Non-cancer<br>women | P value<sup>#</sup> | Adjusted model*<br>Breast cancer<br>survivors | P value<sup>#</sup> | P value<sup>@</sup> |
| --- | --- | --- | --- | --- | --- | --- | --- | --- | --- | --- |
| Insomnia/Sleeplessness |  |  |  |  |  |  |  |  |  |  |
| Never/rarely | 2.14 (0.01) | Ref. | 2.54 (0.06) | Ref. | <0.01 | 2.58 (0.04) | Ref. | 2.62 (0.07) | Ref. | 0.87 |
| Sometime | 2.41 (0.01) | <0.01 | 2.77 (0.03) | <0.01 | <0.01 | 2.66 (0.04) | <0.01 | 2.78 (0.05) | 0.01 | <0.01 |
| Usually | 2.63 (0.01) | <0.01 | 2.75 (0.04) | <0.01 | <0.01 | 2.78 (0.04) | <0.01 | 2.76 (0.05) | 0.05 | 0.80 |
| Daytime Sleepiness |  |  |  |  |  |  |  |  |  |  |
| Never/rarely | 2.35 (0.01) | Ref. | 2.65 (0.03) | Ref. | <0.01 | 2.64 (0.04) | Ref. | 2.69 (0.05) | Ref. | 0.06 |
| Sometime | 2.70 (0.01) | <0.01 | 2.97 (0.05) | <0.01 | <0.01 | 2.75 (0.05) | <0.01 | 2.85 (0.06) | <0.01 | 0.01 |
| Usually | 2.82 (0.03) | <0.01 | 2.98 (0.13) | 0.02 | 0.28 | 2.88 (0.05) | <0.01 | 2.84 (0.13) | 0.19 | 0.75 |

\*. Adjusted by age at recruitment, education, family income, Townsend Deprivation Index, cigarette smoking status, alcohol drinking status, and physical activity levels.
<sup>#</sup>. Within group comparison by levels of insomnia and daytime sleepiness.
<sup>@</sup>. Comparison between breast cancer survivors and non-cancer women

In the adjusted model, after controlling for covariates, all within-group comparisons among controls remained significant at *p* < 0.01. Among breast cancer survivors, those who reported having insomnia “sometime” had significantly higher AL than those reporting no insomnia (*p* ≤ 0.02), but no significant differences were observed between those reporting insomnia ‘usually’ and having insomnia ‘never/rarely’ (*p* = 0.12). A similar pattern was found for daytime sleepiness, such that those reporting having this symptom ‘sometimes’ had significantly higher AL compared to those reporting having no daytime sleepiness (*p* < .01) but differences AL between the ‘never/rarely’ and ‘usually’ groups were not significant (*p* = 0.22). Note, there were no significant differences in AL between those reporting ‘sometime’ and ‘usually’ symptoms for both insomnia and daytime sleepiness, indicating that in both comparisons AL was comparable between the two most severe symptom groups. There were also significant between-group differences in AL between breast cancer survivors and controls who reported having insomnia and daytime sleeplessness ‘sometime’ (*p’s* < .01), indicating that even moderate symptoms of sleep disturbance in breast cancer survivors are associated with higher AL, compared to controls with similar sleep disturbances and survivors without sleep disturbance.

### Predictive Value of Baseline Allostatic Load on Sleep Quality

The pattern of results from the crude model and the adjusted model were nearly identical (Table 2). After adjusting for covariates, despite a general trend of baseline AL being more strongly associated with sleep quality in breast cancer survivors compared to controls, there were no significant group differences in the composite PSQI score or any of the PSQI subdomains at the follow-up assessment. Tests of within-group differences revealed that for both breast cancer survivors and controls sample, AL scores at baseline were negatively associated with sleep quality (PSQI) at follow-up (*p’s* < 0.05). Most subdomains of the PSQI were also negatively associated with baseline AL for both samples (*p* < 0.05) in the adjusted model, with the exceptions of the need for sleep medications for controls and breast cancer survivors, and sleep latency for breast cancer survivors. These results demonstrate a robust negative relationship between baseline AL on sleep quality at follow-up for both cancer survivors and controls.

**Table 2.** Allostatic load at baseline to predict PSQI and its components during the follow-up among breast cancer survivors and controls.

|  | Non-cancer women |  | Breast cancer survivors |  |  |  |
| --- | --- | --- | --- | --- | --- | --- |
| | Crude model<br>Coefficient (95% CI) | Adjusted model<br>Coefficient (95%<br>CI)# | Crude model<br>Coefficient (95% CI) | Adjusted model<br>Coefficient (95% CI)# | Difference<br>$\beta$ (case–ctrl) | Difference<br>(p-value) |
| **PSQI** | 0.244 (0.227, 0.262)* | 0.184 (0.166, 0.202)* | 0.239 (0.130, 0.348)* | 0.207 (0.093, 0.320)* | 0.023 | 0.54 |
| **Sleep duration** | 0.032 (0.028, 0.036)* | 0.020 (0.016, 0.024)* | 0.030 (0.005, 0.055)* | 0.026 (0.001, 0.053)* | 0.006 | 0.55 |
| **Sleep disturbance** | 0.035 (0.033, 0.038)* | 0.038 (0.035, 0.041)* | 0.043 (0.027, 0.059)* | 0.038 (0.021, 0.055)* | 0.000 | 0.99 |
| **Sleep latency** | 0.055 (0.051, 0.060)* | 0.037 (0.032, 0.042)* | 0.038 (0.007, 0.068)* | 0.027 (-0.005, 0.059) | -0.010 | 0.50 |
| **Day dysfunction due to sleepiness** | 0.036 (0.033, 0.039)* | 0.030 (0.027, 0.034)* | 0.034 (0.017, 0.052)* | 0.023 (0.005, 0.042)* | -0.007 | 0.46 |
| **Sleep efficiency** | 0.063 (0.058, 0.069)* | 0.034 (0.028, 0.040)* | 0.064 (0.031, 0.097)* | 0.054 (0.019, 0.089)* | 0.020 | 0.18 |
| **Overall sleep quality** | 0.017 (0.013, 0.20)* | 0.024 (0.020, 0.028)* | 0.028 (0.006, 0.050)* | 0.037 (0.014, 0.060)* | 0.013 | 0.20 |
| **Need med to sleep** | 0.008 (0.005, 0.011)* | 0.001 (-0.003, 0.004) | 0.014 (-0.007, 0.035) | 0.012 (-0.011, 0.034) | 0.011 | 0.13 |

#. Adjusted by age at recruitment, race, education, family income, Townsend Deprivation Index, cigarette smoking status, alcohol drinking status, and physical activity levels.
\*: $p < 0.05$

## Discussion

Understanding the relationship between sleep disturbance and AL in breast cancer survivors may enhance survivorship care by identifying new pathways to reduce the risk of adverse health outcomes. Consistent with our first hypothesis and existing research, the sampled breast cancer survivors had higher AL scores and more severe sleep disturbance symptoms compared to women without cancer. These associations are likely driven by a combination of factors. First, it may be that breast cancer survivors have higher AL even before receiving their cancer diagnosis. Conditions related to key components of AL, such as obesity and metabolic syndrome, have been linked to pathological changes in breast tissue (Naaman et al., 2022) though findings regarding high AL as a risk factor for breast cancer are mixed (Obeng-Gyasi et al., 2024). Early-life stress may confer greater risk for both cancer and high AL later in life via epigenetic mechanisms; while there exists compelling evidence in support of this theory, more research is needed (Mavromanoli et al., 2025). Second, it is likely that receiving a breast cancer diagnosis and subsequent treatment may also cause AL to rise through a combination of stress and treatment side effects (Dieli-Conwright et al., 2016; Nappi et al., 2022). Factors increasing the risk of sleep disturbance among breast cancer survivors include depression, hormonal therapy, chemotherapy, menopause, and pain (Palesh et al., 2013; Leysen et al., 2023). In a large sample, our findings indicate that high AL and sleep disturbance symptoms affect breast cancer survivors at higher rates than peers of similar sex and age, emphasizing the importance of this research and future studies to determine the time course and mechanisms involved.

We found a positive association between AL and sleep disturbance symptom severity among both breast cancer survivors and controls. Although we are cautious to not over interpret the findings in light of their correlational nature, our results suggest that breast cancer survivors and controls demonstrate different patterns of change in AL across levels of sleep disturbance symptom severity. Even after adjusting for covariates, the control sample demonstrated a relatively linear increase in AL across symptom severity strata for both insomnia and daytime sleepiness. This suggests that among adult women with no cancer history, moderate sleep disturbance symptoms are associated with a small but significant increase in AL, while severe sleep disturbance symptoms are associated with a further incremental rise in AL. This same linear relationship between sleep disturbance symptom severity and AL was not found among breast cancer survivors. A significant difference in AL scores was found between survivors with no sleep disturbance symptoms and those with moderate symptoms, but no difference in AL could be detected between those with moderate and frequent symptoms. This pattern may reflect a threshold at which breast cancer survivors begin to evidence high AL, when a moderate level of sleep disturbance is present, with additional sleep disturbance severity not contributing to AL due to a ceiling effect. An important clinical implication is that improving sleep may have significant benefits for breast cancer survivors in reducing AL, even when sleep problems are not a primary concern or do not meet the clinical threshold for a sleep disorder.

Our findings demonstrate that AL at baseline is negatively associated with sleep quality at a later point among both breast cancer survivors and controls, supporting a bidirectional relationship between sleep and AL in combination with our other findings. Most subdomains of the PSQI at follow-up were weakly but significantly associated with baseline AL in both groups. These results suggest that elevated AL may increase the severity of future sleep problems in both breast cancer survivors and the general population, potentially leading to a vicious cycle whereby impaired sleep further worsens AL. This could lead to future downstream effects such as impaired lipid and glucose metabolism (Briançon-Marjollet et al., 2015) and elevated inflammatory markers (Faraut et al., 2012).

While acknowledging the correlational nature of the data, the current findings present potential avenues for future research. Inadequate sleep and high AL are both established risk factors for a range of chronic conditions of public health concern. These include coronary heart disease, stroke, diabetes, depression, anxiety, and cognitive decline (Tu et al., 2024; Clark et al., 2014; Akinyemiju et al., 2020). We have identified breast cancer survivors as a group at high risk for elevated AL and sleep problems in combination, which has significant clinical implications. AL theory suggests that reducing even a single high-risk component or addressing sources of chronic stress can lower overall physiological burden, thereby reducing risk of undesirable health outcomes (Juster et al., 2016). Moreover, evidence of a bidirectional relationship between AL and sleep suggests that improving sleep quality through behavioral intervention is another method of lowering physiological stress. Breast cancer survivors should be informed that even after remission, the risk of poor health outcomes persists. However, medical treatment options, lifestyle changes, and behavioral interventions exist to preempt risk by either targeting components of AL or sleep concerns in breast cancer survivors.

Findings from this study should be interpreted considering several limitations. The components needed to compute allostatic load were only measured in UK BioBank participants during intake. It was therefore not possible to examine change in AL over time, nor was it possible to measure effects of sleep quality on future AL. Because most existing studies examining the relationship between sleep and AL are cross-sectional, and our longitudinal analysis does not include multiple PSQI or AL measurements, the nature of the long-term interaction between sleep and AL should be studied further to better characterize this relationship. Relatedly, baseline sleep assessment relied on single-item measures rather than a widely-used instrument like the PSQI, which was included in only the follow-up wave. While a more comprehensive measurement of sleep quality at all time points would enhance the validity of this analysis, research demonstrates that single-item measures of sleep quality generally meet acceptable reliability and validity standards (Hughes et al., 2017).

Future research should also consider additional psychosocial covariates, such as adverse childhood experiences and recent stressors unrelated to cancer. While race was included as a covariate in our adjusted models, our sample was 95% White and of European descent; representative of the U.K. population from which it was drawn. Results may not generalize to more racially or ethnically diverse populations, particularly as historical marginalization is associated with additional risk for high AL (Van Dyke et al., 2020). Differences in healthcare infrastructure, population demographics, and health outcomes related to poverty mean the findings of this analysis should be applied to U.S. populations with caution. Future research should aim to replicate these findings in more diverse cohorts using repeated PSQI and AL measurements to improve longitudinal insight.

## Conclusion

In summary, this collection of findings indicates a strong relationship between sleep disturbance and AL in both breast cancer survivors and controls that may lead to poor long-term health outcomes. The presence of elevated sleep disturbance and AL among breast cancer survivors, compared to controls, highlights the importance of testing the impact of interventions that address one or both domains in this population to improve long-term health outcomes.

## Data Availability

All data produced in the present work are contained in the manuscript.

## Notes

### Competing Interest Statement

The authors have declared no competing interest.

### Funding Statement

This study did not receive any funding.

### Author Declarations

IRB-Social and Behavioral Sciences (IRB-SBS) of the University of Virginia waived ethical approval for this work.

