## Supplemental Table 1 for "Understanding the relationship between sleep disturbance and allostatic load: A cohort study of women with and without breast cancer"

**Supplemental Table 1**. Selected characteristics in all-female cohort and breast cancer cases

| | Non-cancer women (N=171,938) | Breast cancer survivors (N=5,491) | P value |
|---|---|---|---|
| Age at recruitment, Mean (SD) | 56.35 (7.96) | 59.71 (6.58) | <0.01 |
| Education, N (%) | | | <0.01 |
| High school or less | 62,961 (36.62%) | 2,389 (43.51%) | |
| College / professional | 79,005 (45.95%) | 1,971 (35.90%) | |
| Missing | 29,972 (17.43%) | 1,131 (20.60%) | |
| Family income, N (%) | | | <0.01 |
| < £30,999 | 71,182 (41.40%) | 2,570 (46.80%) | |
| ≥ £30,999 | 71,956 (41.85%) | 1,875 (34.15%) | |
| Missing | 28,800 (16.75%) | 1,046 (19.05%) | |
| Townsend Deprivation Index | | | 0.92 |
| Low | 89,538 (52.08%) | 2,873 (52.32%) | |
| High | 82,223 (47.82%) | 2,612 (47.57%) | |
| Missing | 177 (0.10%) | 6 (0.11%) | |
| Cigarette smoking, N (%) | | | <0.01 |
| Never | 101,405 (58.98%) | 2,992 (54.49%) | |
| Ever | 69,956 (40.69%) | 2,472 (45.02%) | |
| Missing | 577 (0.34%) | 27 (0.49%) | |
| Physical activity | | | <0.01 |
| Low | 24,707 (14.37%) | 844 (15.37%) | |
| Moderate | 57,862 (33.65%) | 1,875 (34.15%) | |
| High | 51,858 (30.16%) | 1,556 (28.34%) | |
| Missing | 37,511 (21.82%) | 1,216 (22.15%) | |
| Alcohol consumption, N (%) | | | <0.01 |
| Special occasions or never | 37,990 (22.10%) | 1,306 (23.78%) | |
| Moderate | 67,997 (39.55%) | 2,086 (37.99%) | |
| Heavy | 65,837 (38.29%) | 2,093 (38.12%) | |
| Missing | 114 (0.07%) | 6 (0.11%) | |
| Insomnia/sleeplessness, N (%) | | | <0.01 |
| Never/rarely | 32,373 (18.83%) | 757 (13.79%) | |
| Sometimes | 845,22 (49.16%) | 2,643 (48.13%) | |
| Usual | 54,973 (31.97%) | 2,087 (38.01%) | |
| Missing | 70 (0.04%) | 4 (0.07%) | |
| Daytime sleepiness, N(%) | | | <0.01 |
| Never/rarely | 135100(78.57%) | 4094(74.56%) | |
| Sometimes | 32331(18.80%) | 1236(22.51%) | |
| Usual | 3937(2.29%) | 148(2.70%) | |
| Missing | 570(0.34%) | 13(0.23%) | |
