## Supplemental Table 2 for "Understanding the relationship between sleep disturbance and allostatic load: A cohort study of women with and without breast cancer"

**Supplemental Table 2:** Distribution of AL scores and AL category in all-female cohort.

| | Control (N=182,028) | Breast cancer Case (N=5,673) | P value |
|---|---|---|---|
| **AL Score** | | | <0.01 |
| **0** | 17214 (9.46%) | 335 (5.91%) | |
| **1** | 44613 (24.51%) | 1162 (20.48%) | |
| **2** | 39843 (21.89%) | 1196 (21.08%) | |
| **3** | 34178 (18.78%) | 1219 (21.49%) | |
| **4** | 24094 (13.24%) | 891 (15.71%) | |
| **5** | 14029 (7.71%) | 546 (9.62%) | |
| **6** | 6092 (3.35%) | 229 (4.04%) | |
| **7** | 1625 (0.89%) | 85 (1.5%) | |
| **8** | 300 (0.16%) | 6 (0.11%) | |
| **9** | 40 (0.02%) | 4 (0.07%) | |
| **10** | 0 | 0 | |
| **11** | 0 | 0 | |
| **AL, continuous (Mean/SD)** | 2.44 (1.63) | 2.74 (1.64) | <0.01 |
