## Supplementary figures and images for "Understanding the relationship between sleep disturbance and allostatic load: A cohort study of women with and without breast cancer"

### Supplemental Figure 3

**Supplemental Figure 1:** Distribution of AL scores in all-female cohort

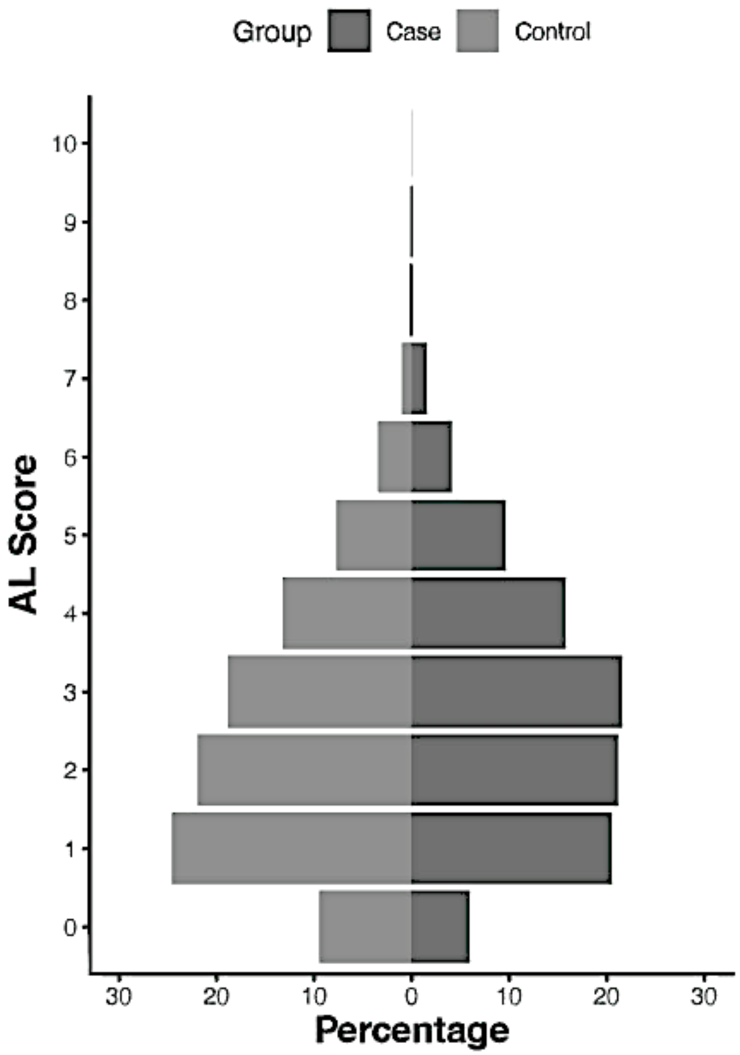
